# Prediction of Venous Thromboembolism Based on Clinical and Genetic Factors

**DOI:** 10.1101/2020.03.05.20031054

**Authors:** David A. Kolin, Scott Kulm, Olivier Elemento

## Abstract

**BACKGROUND:** Both clinical and genetic factors drive the risk of venous thromboembolism. However, whether clinically recorded risk factors and genetic variants can be combined into a clinically applicable predictive score remains unknown.

**METHODS:** Using Cox proportional-hazard models, we analyzed the association of risk factors with the likelihood of venous thromboembolism in U.K. Biobank, a large prospective cohort. We created a novel ten point clinical score using seven established clinical risk factors for venous thromboembolism. We also generated a polygenic risk score of 21 single nucleotide polymorphisms to quantify genetic risk. The genetic score was categorized into high risk (top two deciles of scores), intermediate risk (deciles three to eight), and low risk (lowest two deciles). The discrete clinical score led to the following approximate decile categorizations: high risk (5 to 10 points), intermediate risk (3 to 4 points), and low risk (0 to 2 points).

**RESULTS:** Amongst the 502,536 participants in the U.K. Biobank, there were 4,843 events of venous thromboembolism. Analyses of established clinical risk factors and the most commonly used medications revealed that participants were at decreased risk of venous thromboembolism if they had ever used oral contraceptive pills (hazard ratio, 0.88; 95% confidence interval [CI], 0.79 to 0.99) or if they currently used bendroflumethiazide (hazard ratio, 0.84; 95% CI, 0.74 to 0.95), cod liver oil capsules (hazard ratio, 0.87; 95% CI, 0.77 to 0.99), or atenolol (hazard ratio, 0.79; 95% CI, 0.68 to 0.91). Participants were at significantly increased risk of venous thromboembolism if they were at high clinical risk (hazard ratio, 5.98; 95% CI, 5.43 to 6.59) or high genetic risk (hazard ratio, 2.28; 95% CI, 2.07 to 2.51) relative to participants at low clinical or genetic risk, respectively. Combining clinical risk factors with genetic risk factors produced a model that better predicted risk of venous thromboembolism than either model alone (P<0.001). Participants at high clinical and genetic risk in the combined score had over an eightfold increased risk of venous thromboembolism relative to participants at low risk (hazard ratio, 8.27; 95% CI 7.59 to 9.00).

**CONCLUSIONS:** By assessing venous thromboembolic events in over 500,000 participants, we identified several known and novel associations between risk factors and venous thromboembolism. Participants in the high risk group of a combined score, consisting of clinical and genetic factors, were over eight times more likely to experience venous thromboembolism than participants in the low risk group.

## INTRODUCTION

Both clinical and genetic factors drive the likelihood of venous thromboembolism, the leading cause of preventable hospital deaths.^1^ Considerable evidence demonstrates that individuals exposed to clinical risk factors, such as cigarette smoke, cancer, oral contraceptive pills, high body mass index, recent hospitalization, and major surgery, have markedly increased risk of venous thromboembolism.^2–7^

While clinical factors account for a significant proportion of thromboembolic risk, over 60% of variation in the risk of venous thromboembolism can be attributed to genetic factors.^8^ Factor V Leiden and Factor II Mutation, two monogenic variants, were first described in the 1990s.^9–14^ Since then, genome-wide association studies have identified over 20 additional loci.^15–19^ Risk alleles, when combined into a polygenic score, are capable of quantifying genetic susceptibility and are often more effective at predicting risk than rare monogenic variants alone.^20–27^

Currently, our understanding of clinical and genetic risk for venous thromboembolism guides both prophylaxis and treatment.^28–31^ In acute care settings, clinical scores, such as the Wells Score and Geneva Score, can rapidly quantify risk of venous thromboembolism. Genetic susceptibility to venous thromboembolism can be ascertained by detecting specific point mutations known to cause inherited thrombophilias. However, a quantitative scoring system – consisting of clinical and genetic factors – that predicts the likelihood of venous thromboembolic events prior to hospitalization has not yet been established. In order to address these gaps, we first explored clinical risk factors for venous thromboembolism in U.K. Biobank, a large longitudinal cohort. We then determined the extent to which clinical and genetic factors, both individually and combined, predispose individuals to venous thromboembolism.

## METHODS

### Study Population

The U.K. Biobank is a prospective cohort of 502,536 participants between the ages of 40 and 69 years recruited from across the United Kingdom from 2006 to 2010. Baseline information was gathered during an in-person interview, and in-patient health outcome data were collected prospectively for all hospital episodes. For this study, all U.K. Biobank participants were included in analyses of clinical risk factors. For the polygenic risk score analyses, 131,194 participants were included in score derivation, and 276,988 participants were included in model assessment. Written informed consent was obtained for all of the participants in U.K. Biobank.

### Study Outcome

The study outcome was any primary venous thromboembolism event, including portal vein thrombosis, thrombophlebitis migrans, pulmonary embolism, combined phlebitis thrombophlebitis, and embolism and thrombosis of other veins. Additional details regarding the coding of venous thromboembolism events are provided in Table S1 in the Supplementary Appendix, available with the full text of this article at NEJM.org. Participants were considered at risk for venous thromboembolism at baseline and were censored at death, loss to follow-up, or the last date of hospital admission (March 31, 2017 for England, October 31, 2016 for Scotland, and February 29, 2016 for Wales).

### Polygenic Risk Score

We constructed a polygenic risk score from 21 single nucleotide polymorphisms (SNPs), all with genome-wide significance in previously published genome wide association studies of venous thromboembolism.^15,16,18,19,32^ To determine which studies and SNPs to include in the polygenic score, a forward stepwise selection method, considering both dominance effects and genetic interactions, was developed. Variable strength amongst studies was measured by the effect sizes, generated from a logistic regression of all study specific scores. The polygenic score was then constructed from the selected SNPs by summing the product of each participant’s number of alleles, the study-derived effect size, and a derived multiplier representing the strength of each study. Logistic regression analyses employed in the derivation of the polygenic risk score were adjusted for basic covariates: the first four principal components of ancestry, the genotyping array, age, and sex. Details of the included studies and the genotyping platform used are provided in Table S2 in the Supplementary Appendix.

### Statistical Analysis

We tested the association between eleven established risk factors and incident venous thromboembolism events using Cox proportional-hazard models. In order to better understand the association of potential risk factors with venous thromboembolism, we also conducted four additional analyses of the ten most common cancer subtypes, medications, non-cancer illnesses, and fracture sites using Cox proportional-hazard models. Primary Cox proportional-hazard models were adjusted for age, sex, body mass index (BMI), previous cancer diagnosis, smoking status, alcohol intake frequency, use of oral contraceptive pills, use of hormone replacement therapy, fracture in the last five years, previous deep vein thrombosis, previous pulmonary embolism, and the first four principal components of ancestry.

Next, we created three novel scores for venous thromboembolism: a clinical risk score, a genetic risk score (based on polygenic risk), and a combined score. We created a ten point clinical risk score using seven established risk factors for venous thromboembolism: sex, age, BMI, smoking status, fracture in the last five years, previous cancer diagnosis, and previous venous thromboembolism (deep vein thrombosis or pulmonary embolism) (Table S3 in the Supplementary Appendix). After creating a genetic risk score based on polygenic risk, the clinical and genetic scores were combined into a single score by adding each score proportional to their hazard ratios, derived from a training set.

The genetic and combined score were then used to categorize study participants into three risk categories: low risk (lowest two deciles), intermediate risk (deciles three to eight), and high risk (top two deciles). The discrete ten point clinical score led to approximate decile categorizations as follows: high risk (5 to 10 points), intermediate risk (3 to 4 points), and low risk (0 to 2 points) (Table S4 in the Supplementary Appendix). These categories were used with Cox proportional-hazard models to calculate hazard ratios, 10-year event rates, and Kaplan-Meier estimators. The fit of each model was measured by using concordance for Cox proportional-hazard models and area under the curve (AUC) for logistic regression models. All of the analyses were performed with the use of R software, version 3.5 (R Project for Statistical Computing).

## RESULTS

### Participant Characteristics and Established Risk Factors

There were a total of 4,843 venous thromboembolic events amongst the 502,536 participants in U.K. Biobank. The mean age of participants was 56.5 years, and 54.4% of participants were female (Table S5 in the Supplementary Appendix). At baseline, 220,446 (80.6%) women reported ever taking an oral contraceptive pill. 9,323 (1.8%) participants had a previous deep vein thrombosis, and 3,955 (0.8%) participants had a previous pulmonary embolism.

We examined 11 known risk factors for venous thromboembolism using multivariate Cox-proportional hazard models. Class 3 obesity (BMI ≥ 40 kg/m^2^), relative to normal weight, was one of two risk factors associated with over a threefold increase in the risk of venous thromboembolism (hazard ratio, 3.30; 95% confidence interval [CI], 2.81 to 3.88) (Fig. 1). Previous deep vein thrombosis was the only other risk factor associated with over a threefold increase in risk (hazard ratio, 3.34; 95% CI, 2.96 to 3.77). Perhaps surprisingly, participants who had ever used oral contraception had a 12% decreased risk of venous thromboembolism (hazard ratio, 0.88; 95% CI, 0.79 to 0.99). In exploratory analyses of the length of use of contraception, we found that women who used contraception for at least twenty years were also at decreased risk of venous thromboembolism (hazard ratio, 0.81; 95% CI, 0.67 to 0.98) (Table S6 in the Supplementary Appendix). However, an insignificant trend of venous thromboembolism risk was observed when length of oral contraceptive use was analyzed on a continuous scale (P=0.29). Further analyses revealed that, of the 4,919 current users of oral contraceptive pills, 935 were taking desogestrel (Cerazette 75 microgram tablets) (Table S7 in the Supplementary Appendix). Compared to fully adjusted Cox-proportional hazard models, univariate risk ratios of death from venous thromboembolism identified similar patterns in risk (Fig. S1 and Table S8 in the Supplementary Appendix).

**Figure 1.**
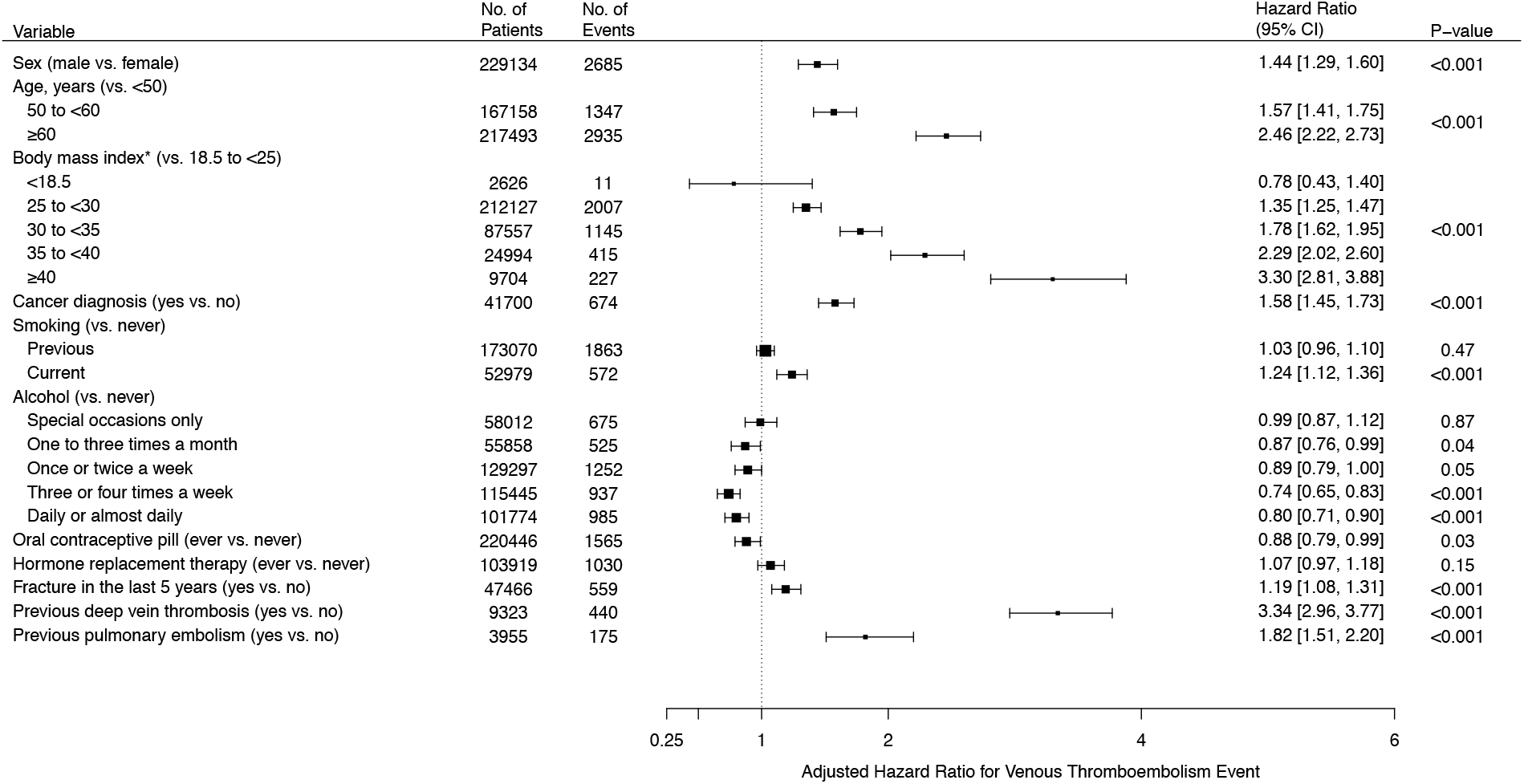
Adjusted Hazard Ratios for Venous Thromboembolism for Established Risk Factors. Shown are the adjusted hazard ratios for venous thromboembolism for eleven established risk factors. For any individual risk factor, Cox regression models were adjusted for all other established risk factors and the first four principal components of ancestry. The I bars represent 95% confidence intervals. Body mass index (*) was measured in kg/m^2^.

### Cancer Subtypes, Medications, Non-Cancer Illnesses, and Fracture Sites

In order to better understand the specific clinical risk factors associated with risk of venous thromboembolism, we also analyzed the association of common cancer subtypes, medications, non-cancer illnesses, and fracture sites with venous thromboembolic risk. Previously diagnosed breast cancer, prostate cancer, colon cancer, and cervical cancer were associated with increased risk of venous thromboembolism (Fig. 2). Of the ten most common medications in U.K. Biobank, three were associated with a decreased risk of venous thromboembolism and one was associated with increased risk. Hazard ratios for bendroflumethiazide and atenolol were 0.84 (95% CI, 0.74 to 0.95) and 0.79 (95% CI, 0.68 to 0.91), respectively, and there was some evidence that use of cod liver oil capsules was associated with a decreased risk of venous thromboembolism (hazard ratio, 0.87; 95% CI, 0.77 to 0.99). Amitriptyline was the only medication associated with an increased risk of venous thromboembolism (hazard ratio, 1.23; 95% CI, 1.02 to 1.48). Participants with asthma were at 16% increased risk of venous thromboembolism, while those with either osteoarthritis or depression were at 18% increased risk of experiencing a venous thromboembolic event. Hypercholesterolemia was associated with an 8% decreased risk of venous thromboembolism (hazard ratio, 0.92; 95% CI, 0.84 to 1.00), and diabetes was associated with a 20% decreased risk of venous thromboembolism (hazard ratio, 0.80; 95% CI, 0.70 to 0.92). However, evidence of any association disappeared with adjustment for common therapies for hypercholesterolemia or diabetes (Table S9 in the Supplementary Appendix). Of the participants who fractured a bone in the last five years, only participants who fractured their wrist or unspecified bones had significantly increased risk of venous thromboembolic events. We found that analyses with minimally adjusted models generated similar results (Fig. S2 through S6 in the Supplementary Appendix).

**Figure 2.**
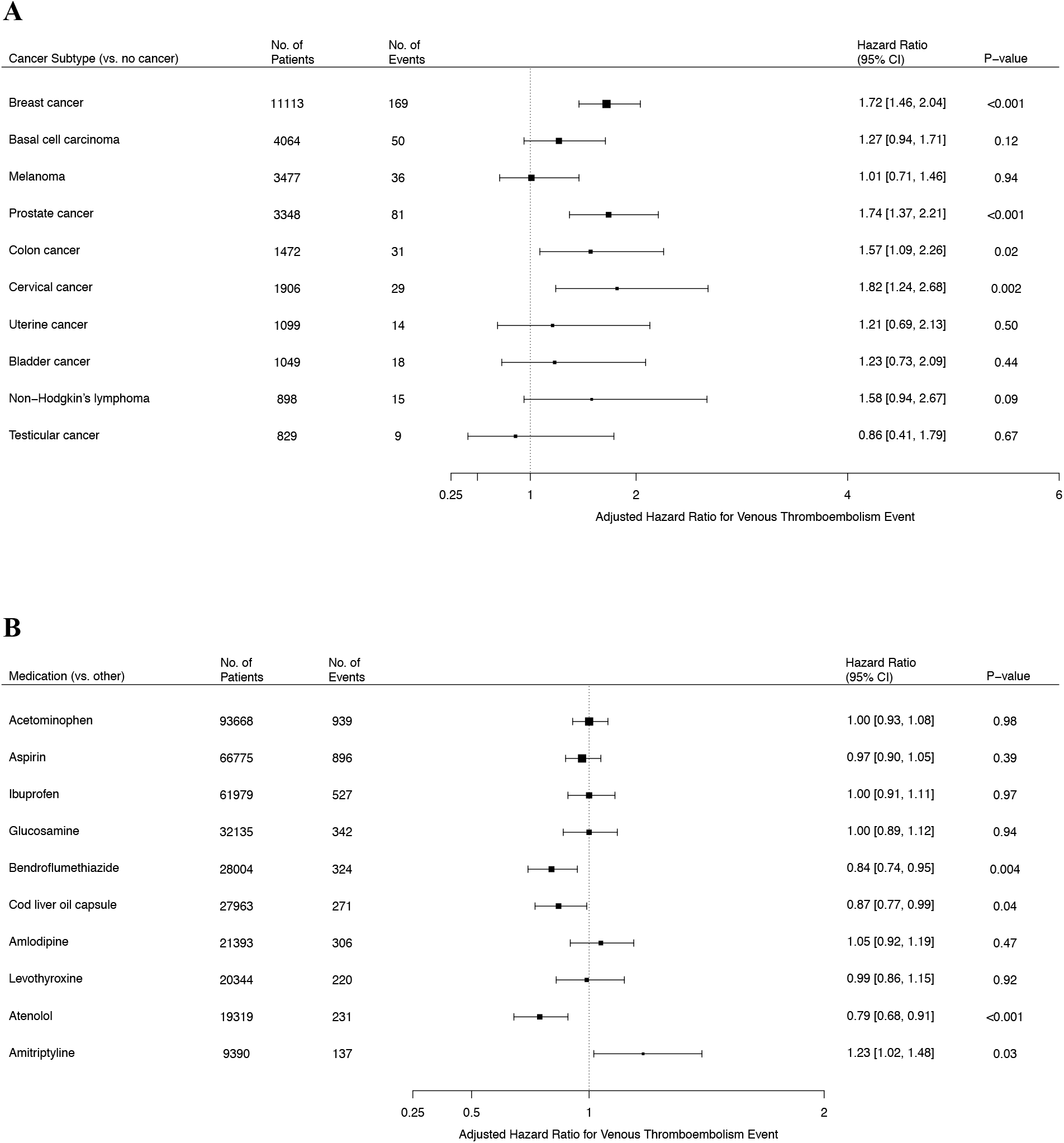
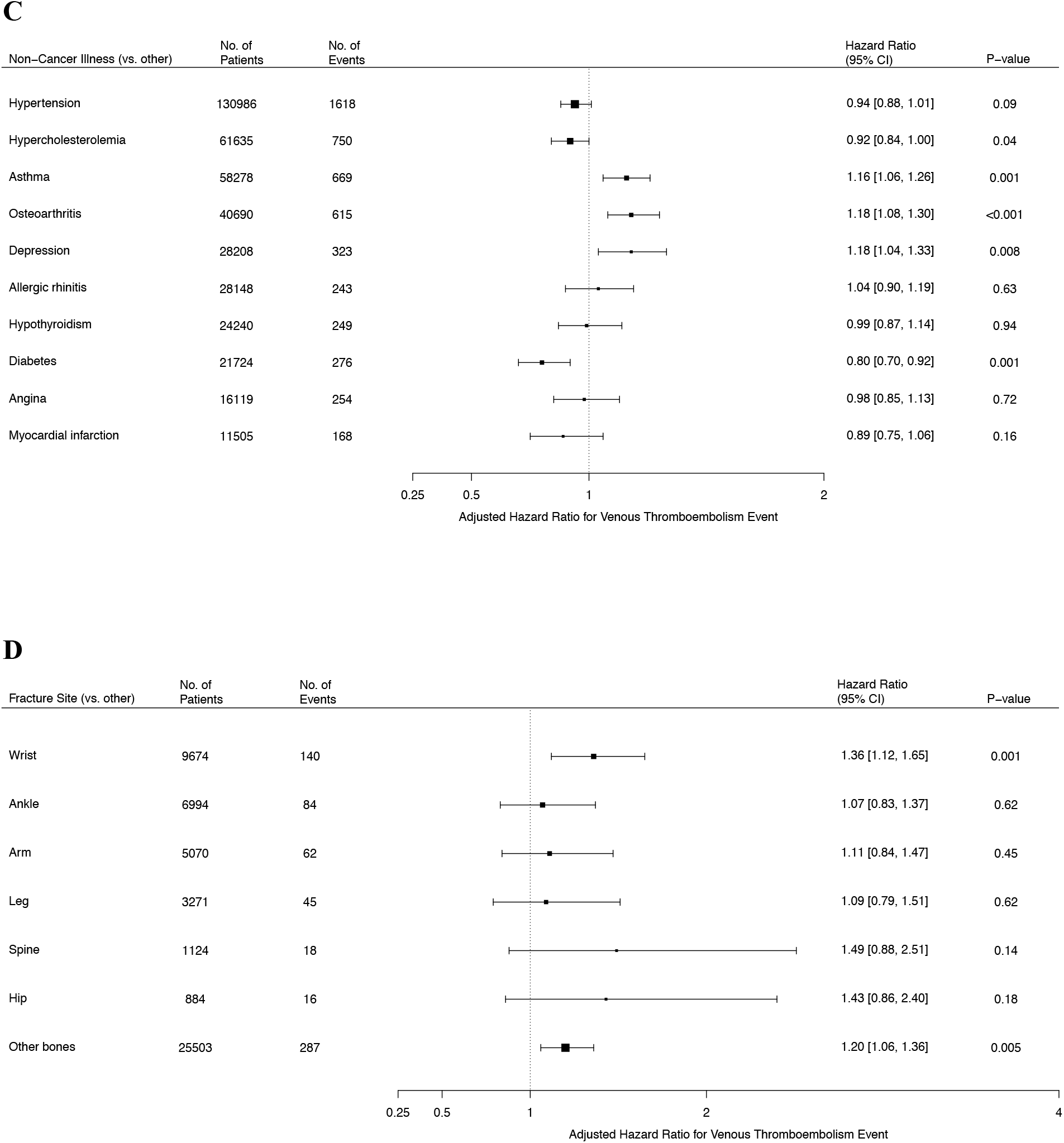
Adjusted Hazard Ratios for Venous Thromboembolism for Common Cancer Subtypes, Medications, Non-Cancer Illnesses, and Fracture Sites. Shown are the adjusted hazard ratios for venous thromboembolic events for four factors of interest: common cancer subtypes, medications, non-cancer illnesses, and fracture sites. All estimates were adjusted for all eleven established risk factors and the first four principal components of ancestry. The I bars represent 95% confidence intervals

### Model Creation

We compared nested Cox proportional-hazard and logistic regression models of the clinical, genetic, and combined scores in order to determine whether the addition of covariates improved the predictive abilities of our models. The Cox proportional-hazard model adjusted for basic covariates alone generated a concordance of 0.63 (95% CI, 0.62 to 0.64) (Figure 3). A model with the clinical risk score, adjusted for the first four principal components of ancestry and genotyping array, improved concordance to 0.68 (95% CI, 0.66 to 0.69). Similarly, a model with the genetic score, adjusted for the basic covariates, improved concordance relative to the basic model alone (concordance, 0.67; 95% CI, 0.66 to 0.69). The odds ratio for participants in the top polygenic risk score percentile compared to the bottom 99 percentiles was 5.33 (95% CI, 5.32 to 5.34), and polygenic risk increased significantly with the number of venous thromboembolic events (Fig. S7 and S8 in the Supplementary Appendix). The combined score, adjusted for the first four principal components of ancestry and the sequencing array, generated a concordance of 0.71 (95% CI, 0.70 to 0.72), significantly higher than either previous model alone (P<0.001). Equivalent analyses with logistic regression generated an AUC of 0.70 (95% CI, 0.69 to 0.71). Cross validation of logistic regressions and bootstrap analyses of the Cox proportional-hazard models led to comparable results. Additional analyses showed an absence of additional interactions or clustering (Fig. S9 to S13 in the Supplementary Appendix).

**Figure 3.**
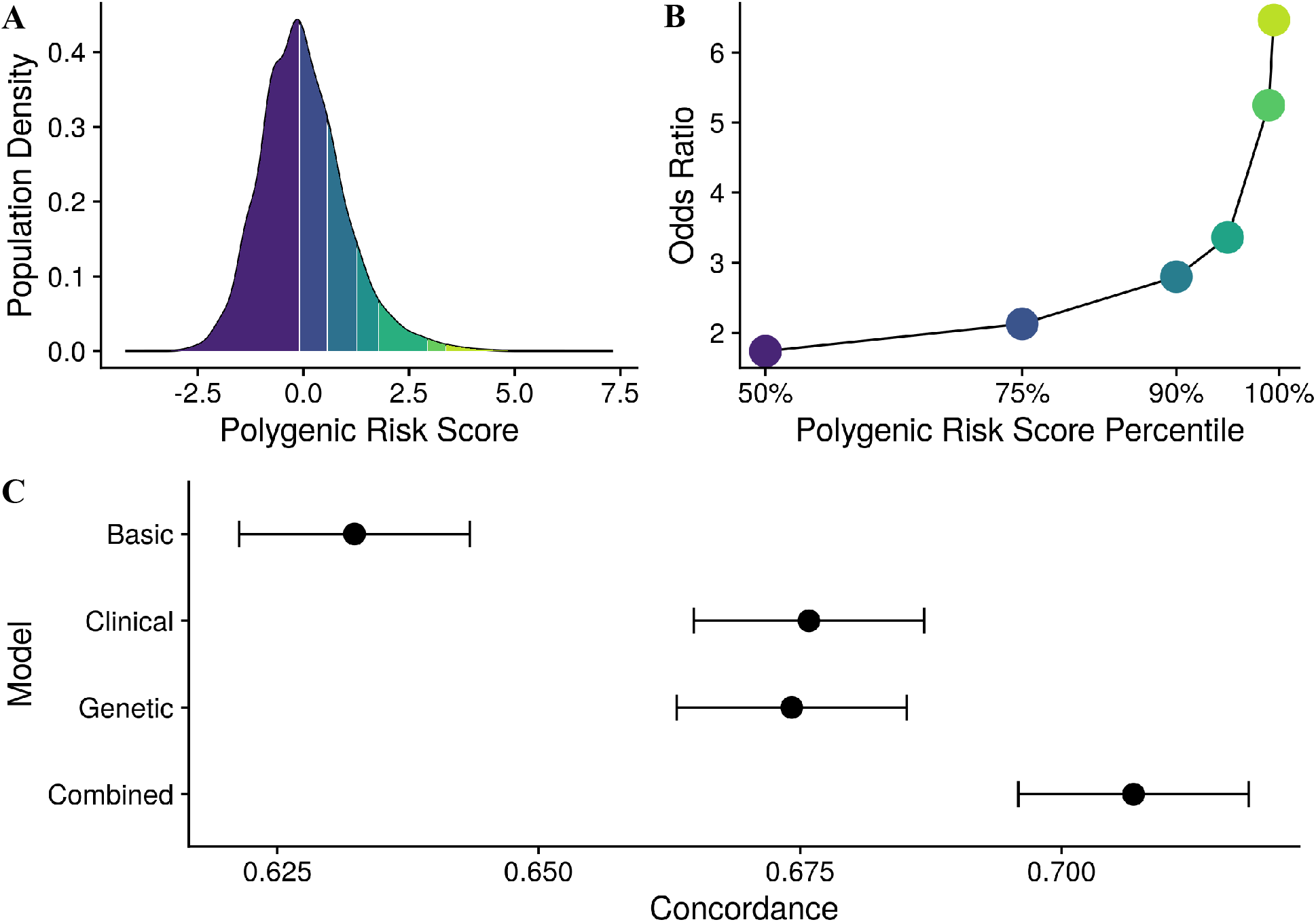
Model Performance of a Basic, Clinical, Genetic, and Combined Score. Panel A shows the density of the polygenic risk score stratified by the 50^th^, 75^th^, 90^th^, 95^th^, 99^th^ and 99.5^th^ percentiles. Panel B shows the odds ratio for each polygenic risk score percentile. Panel C shows the concordance values, derived from adjusted Cox-proportional hazard models, for the basic, clinical, genetic, and combined scores. The concordance of the combined model is significantly higher than the concordance of each of the other models (P<0.001). The I bars represent 95% confidence intervals.

### Risk Stratification

Clinical, genetic and combined scores were used to stratify risk by comparing participants at low, intermediate, and high risk. Participants at high clinical or high genetic risk had 498% (hazard ratio, 5.98; 95% CI 5.43 to 6.59) and 128% (hazard ratio, 2.28; 95% CI 2.07 to 2.51) increased risk of venous thromboembolism, respectively, relative to participants at low clinical or low genetic risk. Participants at high genetic risk yet low clinical risk had an 80% (hazard ratio, 0.20; 95% CI 0.17 to 0.23) decreased risk of venous thromboembolism compared to participants at high genetic and high clinical risk, suggesting that clinical factors can attenuate genetic predispositions. Alternatively, participants at high clinical and genetic risk had over a twofold (hazard ratio, 2.09; 95% CI, 1.66 to 2.65) increased risk of venous thromboembolism compared to participants at high clinical yet low genetic risk, indicating the added benefits of accounting for genetic factors. For the combined score, participants in the high risk group were at over an eightfold (hazard ratio, 8.27; 95% CI 7.59 to 9.00) increased risk of venous thromboembolism relative to participants in the low risk group (Fig. S14 in the Supplementary Appendix). When the clinical score and genetic score were analyzed independently, participants at both high clinical and genetic risk, 2.7% of the total population, had over thirteen-fold (hazard ratio, 13.87; 95% CI 11.22 to 17.13) greater risk of venous thromboembolism than participants at both low clinical and genetic risk (Figure 4). The 10-year event rate was 4.89% for participants at high clinical and genetic risk, and 0.37% for participants at low clinical and genetic risk.

**Figure 4.**
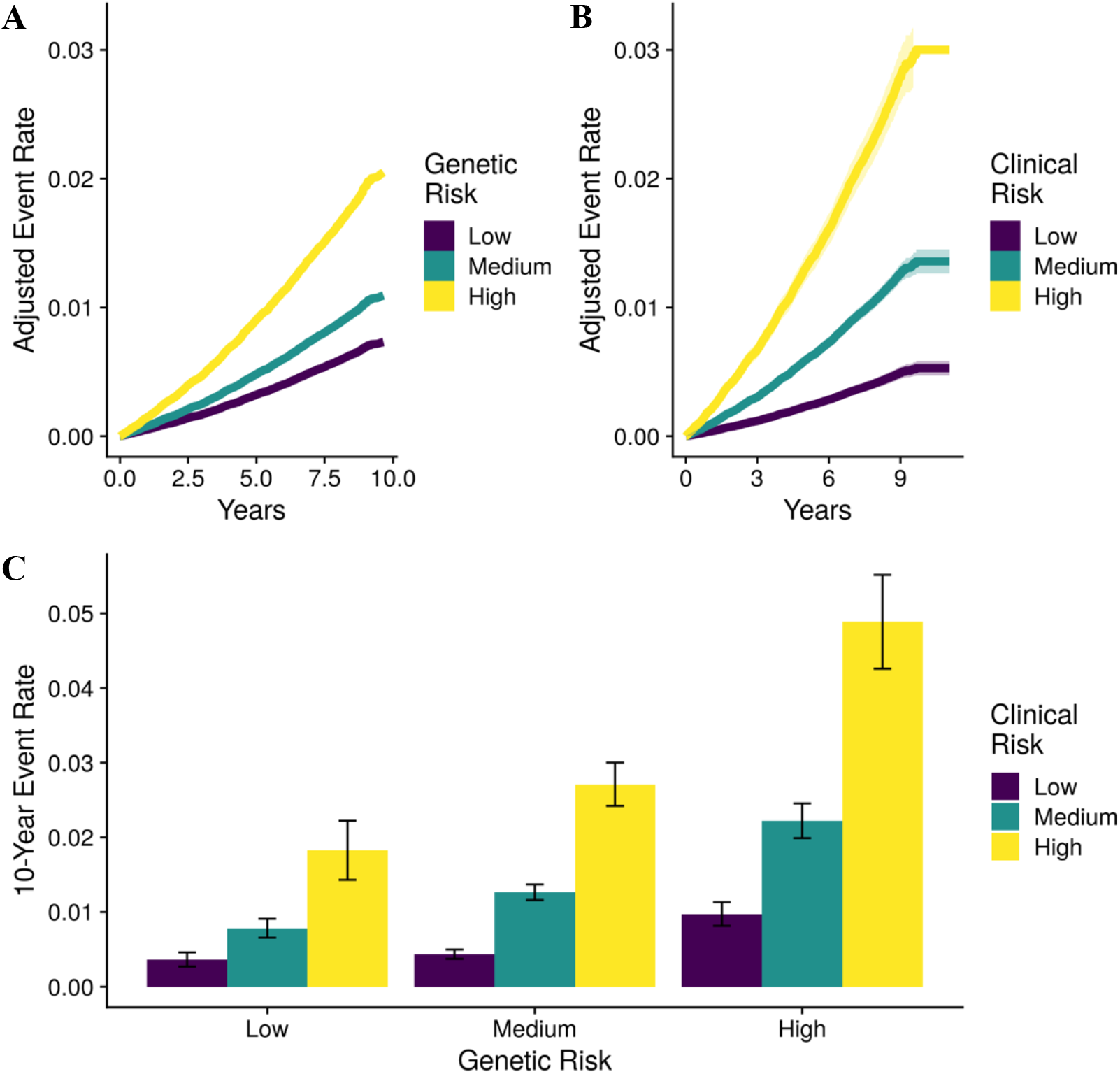
Prediction of Venous Thromboembolism. Panel A and B show the adjusted Kaplan-Meier curves for the genetic and clinical score, respectively. Panel C shows 10-year event rates for venous thromboembolism, stratified by both the clinical score and genetic score. The I bars represent 95% confidence intervals.

## DISCUSSION

In this study, we quantified the risk of venous thromboembolism by examining both clinical and genetic risk factors in U.K. Biobank, a large prospective cohort of over 500,000 participants. We found several noteworthy associations, and we subsequently derived a novel risk score that combines both clinical and genetic factors. Our findings support four conclusions.

First, in analyses of clinical risk factors, we found that participants who had ever used oral contraceptives were at decreased risk of venous thromboembolism (hazard ratio, 0.88). This finding is in direct contrast to the well-described increased risk of venous thromboembolism with oral contraceptive use.^33^ The reasons for this finding are likely multifold. Importantly, while the majority of women had reported ever using oral contraceptive pills, most women had discontinued use by the start of the study. Furthermore, the mean age of participants in our cohort at baseline was over 56 years, suggesting that women who had ever used oral contraceptive pills were decades past the high-risk period for venous thromboembolism that occurs during the first months of oral contraceptive use. Finally, due to contraindications, participants with severe thrombophilias likely did not use oral contraceptive pills.

Second, analyses of the most common cancer subtypes, medications, non-cancer illnesses, and fracture sites identified several associations. The decreased risk of venous thromboembolism with use of atenolol may be due to lower levels of circulating coagulation factors.^34^ While the reduced risk of venous thromboembolism with use of cod liver oil capsules has been reported previously, to the best of our knowledge, use of bendroflumethiazide has not been associated with a decreased risk of venous thromboembolism.^35,36^ While it is plausible that glucocorticoid and antidepressant use contribute to our findings that asthma, osteoarthritis, and depression are associated with increased risk of venous thromboembolism, accounting for common disease-specific medications does not reduce the likelihood of venous thromboembolism considerably. Alternatively, we provide evidence that the apparent protective effects of hypercholesterolemia and diabetes in this cohort are likely due to use of disease-specific therapies.

Third, combining clinical and genetic factors into a single combined score yielded a predictive model with accuracy similar to that of other scoring systems currently used in clinical settings (AUC, 0.70; concordance, 0.71). For example, the Wells Score and Geneva Score, two scores frequently used to assess short-term risk of pulmonary embolism, have reported AUCs ranging from approximately 0.68 to 0.85 and 0.64 to 0.76, respectively.^37–40^ While both the Wells Score and Geneva Score evaluate risk in hospitalized patients, the combined clinical and genetic score presented herein provides a means to estimate risk of venous thromboembolism in individuals who are not hospitalized.

Fourth, the ability of a clinical risk score for venous thromboembolism to stratify participants into different risk groups is improved by the addition of a polygenic risk score. Even after participants are categorized according to their clinical risk, by using seven major risk factors for venous thromboembolism, a polygenic risk score allows for further categorization of participants into significantly different risk groups. These data indicate that genetic risk factors are powerful modulators of susceptibility to venous thromboembolism, and our results suggest that adoption of polygenic risk scores in the clinic may improve venous thromboembolism prediction and prophylaxis.

Our study has several limitations. First, although we attempted to control for confounding through multivariate modeling including fifteen covariates, residual confounding still remains, from variables such as immobility and diet. Second, this study relied on a single cohort of primarily Caucasian participants, and the relative homogeneity of U.K. Biobank – derived from a population of participants from the United Kingdom – may have resulted in models that overestimate the true predictive power of our scoring system. Third, the duration of this prospective study was slightly greater than ten years, a time frame which may underestimate the true predictive power of the clinical and genetic risk scores.

In conclusion, analysis of thromboembolic events in over 500,000 participants identified several known and novel associations. Furthermore, synergistically combining genetic and clinical risk factors into a single combined score identified that participants in the top two deciles of the score were at over eightfold increased risk of venous thromboembolism relative to participants in the lowest two deciles.

## Data Availability

Any data is available upon request.

